# Basal cell carcinoma treated with HeberFERON. A real world retrospective study

**DOI:** 10.1101/2021.09.24.21263710

**Authors:** C Martínez-Suárez, Y Roben-Aguilar, O Reyes-Acosta, Y Garcia-Vega, J Vega-Abascal, V Sánchez-Linares, D Sotolongo-Díaz, Y Piña-Rodriguez, M Fernández-Martori, A Betancourt-Pérez, M Jimenez-Lamas, Y Ballester-Caballero, R Pérez-Morgado, M Curbelo-Alonso, A Molina-Abad, R Martínez-Borrego, J Maturell-Peraza, L Pulido-Garcia, N López-Pupo, Y Ramírez-Hidalgo, M Ramos-Trujillo, I Fernández-Ramirez, M Hernández-Colina, A Perez-Lopez, Y Leon-Garcia, S Chaya-Salgado, Y La O-Ayala, R Hernández-Rodriguez, Y Duncan-Roberts, I Bello-Rivero

**Affiliations:** Department of Clinical Investigations, Center for Genetic Engineering and Biotechnology (CIGB); Production Direction, Center for Genetic Engineering and Biotechnology (CIGB); Promotion and Distribution Direction, Center for Genetic Engineering and Biotechnology (CIGB); Department of Clinical Trial, Center of Molecular Immunology (CIM), Havana; Jose Avila Serrano Polyclinic, Holguin; Center Polyclinic, Santi Spiritus; Department of Dermatology, Antonio Luaces Iraola Hospital, Ciego de Avila; Department of Maxillofacial Surgery, Antonio Luaces Iraola Hospital, Ciego de Avila; Faustino Perez Hospital, Colon, Matanzas; Carlos J. Finlay Polyclinic, Colon, Matanzas; Arnaldo Milian Castro Hospital, Villa Clara; Maria Curi Oncological Hospital, Camagüey; Amalia Simoni Hospital, Camagüey; Gustavo Aldereguia Hospital, Cienfuegos; Pablo Noriega Polyclinic, Mayabeque; Comandante Pinares Hospital, San Cristobal, Artemisa; Heroes del Baire Hospital, Island of Youth; Celia Sanchez Manduley Hospital, Manzanillo, Granma; Juan Bruno Zayas Hospital, Santiago de Cuba; Ernesto Guevara Hospital, Las Tunas; Leon Cuervo Rubio Hospital, Pinar del Rio; Agustinho Neto Hospital, Guantánamo; Havana University; Calixto Garcia Hospital; Center of Medical Surgical Investigations (CIMEQ); Salvador Allende Hospital; Manuel Fajardo Hospital, Havana. Cuba

**Keywords:** basal cell carcinoma, multiple basal cell carcinomas, interferon combination

## Abstract

**Background:** Basal cell carcinoma is the most common type of skin cancer with major impact in health-related quality of life. The use of the formulation based on the combination of IFN-alpha 2b and IFN-gamma (HeberFERON) is an effective alternative in the treatment of basal cell carcinoma, immunogenic tumor, potentially responsible to immunotherapies. The aim of this report is to record, retrospectively, the effect of HeberFERON patients with BCC in the Cuban real word condition.

**Methods:** This is a retrospectively study of the use of HeberFERON in real world conditions. Eligible patients were adults with histologic diagnosis of single or multiple basal cell carcinoma of any skin phototype, lesions of any size, subtype, location, recurrent or not, with or without specific prior treatments. Adult patients, who signed the informed consent to receive the treatment with HeberFERON, were identified from the data bases. The evaluation of clinical effectiveness was carried out according to RECIST 1.1. Ethical committee of participating institutions approved the study.

**Results:** In clinical practice evaluated patients the nose was the region of higher frequency of tumors (36.3%) and the nodular clinical subtype was the predominant (45.3%). Clinical response rate differences (p=0. 000) were found, with complete response of 61.9%, and partial response of 32.7%; with an overall response rate of 94.2% The HeberFERON exerted a 100% disease control, with no progression reported in 640 treated patients. The best responder tumor subtypes to HeberFERON were the more aggressive tumors, morpheaform with complete response of 72% (overall response=96%), followed by the infiltrative with complete response of 66.7% (overall response=100%). Tumor with larger size and patients with more than four tumors had lesser response to the anti-tumor effect of HeberFERON.

**Conclusions:** HeberFERON was highly effective in basal cell carcinomas in real world conditions. In the context of resistance of skin tumors to hedgehog and immune check point inhibitors the combination of IFNs alpha 2b and IFN gamma appears as a plausible therapeutic option for a wide number of basal cell carcinomas.

## Introduction

Basal cell carcinoma (BCC) is the most common type of skin cancer worldwide^1^ and the incidence rates rise continuously^2^. Cuba is currently one of the Latin American and Third World countries with the highest incidence of non-melanoma skin cancer with > 6000 male and > 5000 female cases diagnosed annually, representing a rate of 152.8 and 126.8 per 100,000 inhabitants, respectively^3^.

BCC is a neoplasm with low metastatic potential and slow growth, but locally invasive and destructive. It affects the head and neck region in approximately 70% of cases. Patients diagnosed with a first BCC are at increased risk of developing a second BCC. Simultaneous or consecutive development of new BCCs has a negative impact on clinical outcome, increasing morbidity, and requiring repeated therapeutic interventions that imposes a greater healthcare cost burden^4^. In addition, patients with a history of BCCs also have an elevated risk of developing other cancers related to ultraviolet radiation^5,6^. The prognosis for patients who receive appropriate therapy is habitually good^7^.

Several clinical subtypes of BCC have been described in the literature. The main clinical subtypes are nodular, superficial, and morpheaform BCC^8^. Nodular BCC is the most common clinical subtype, accounting for 50–79% of all BCCs. Superficial BCC is the second most common clinical subtype of BCC, representing 15% of BCC ^5,9^, morpheaform BCC is the 10% of BCC, whereas only 1% of BCC evolves in a giant BCC or in ulcus rodens^10^.

Risk factors for the development of BCC include fair skin type, exposure to ultraviolet radiation or radiation therapy, long-term exposure to arsenic, age, gender, history of BCC, genetic disorders (eg, Gorlin syndrome, xeroderma pigmentosum), and immunosuppression^11,12,13,14^.

Prognostic factors for BCC has been described, and included tumor size (larger size leads to a higher risk of recurrence), clinical margins (poorly defined lesions are at higher risk), histological subtype (morpheaform, and metatypical BCC represent high risk lesions), histological features (perineural and/or perivascular invasion are markers of higher risk), recurrence, and tumor location^15^.

For small tumors or intermediate-sized, the treatment of choice is surgical excision, whereas Mohs micrographic surgery is preferentially indicated for higher-risk or recurrent tumors, tumors in specific anatomic locations, and those with a wider diameter^16,15,17^. Multiple, larger tumors and more complex locations constitute a therapeutic challenges. Vismodegib and Sonidegib, inhibitors of hedgehog signaling are oral treatments of BCC, when surgery or radiotherapy is not appropriate^18^; however, mechanisms of resistance to these inhibitors have been identified^19^. For these reasons, it is convenient to have new treatment alternatives that have specific advantages over current ones, that offer possibilities for non-responders and relapses.

The use of a formulation based on the combination of IFN-alpha 2b and IFN-gamma is an effective alternative in the treatment of malignant diseases, with increased biological potency sustained in the synergistic anti-proliferative effect^20,21^. After the approval of HeberFERON^®^ for BCC in Cuba in 2016, it was introduced widely in medical practice for the treatment of patients with BCC at risk of recurrence or mutilation, advanced BCC or multiple BCC. The aim of this report is to record, retrospectively, the effect of HeberFERON^®^ in these cohorts of patients in the Cuban real word condition.

## Methods

### Study design and patients

This was a retrospective study of patients treated with HeberFERON (CIGB, Havana; approved, for clinical use in BCC of any subtype, localization and size by Cuban Regulatory Authority in August 04, 2016),^22^ in the context of the National Cuban Extension Program with the use of HeberFERON® (NCEP-HFN).

The study universe was made up of all adult patients, residing in Cuba, of any gender and skin color with a histologic diagnosis of BCC who attended the dermatology, maxillofacial or superficial tumors consultations of the hospitals or polyclinics that participated in the study. Eligible patients were adults (age ≥19 years) with single or multiple BCCs of any size, subtype, location, recurrent or not, with or without tumor specific prior treatments. A punch biopsy of not more than 25% of the total lesion size confirmed the diagnosis of BCC, a requisite to be treated with HeberFERON^®^.

Adult patients, who signed the informed consent to receive the treatment with HeberFERON^®^ and the use of personal data and laboratory results, were identified from the data base from departments of dermatology, maxillofacial, or peripheral tumors from several secondary or primary care institutions in Cuba. Ethical oversight of the study was waived by ethics and research institutional committees of participating institutions the study.

The study was done in 14 provinces and the special municipality of Island of Youth, in corresponding hospitals and policlinics of Cuba. Each patient had a medical history, a physical examination, a skin examination for disease assessment, documentation of prior treatments, or medications, and laboratory tests (hemoglobin determination and platelets and white blood cell total and differential counts).

Control variables were taken into account such as: age, skin color, gender, stage of the disease, initial size of the lesion, number of lesions, tumor type (primary, recurrent), clinical subtype, location of the lesion and skin phototypes according to Fitzpatrick^23^. The evaluation of clinical effectiveness was carried out according to RECIST 1.1^24^.

### Procedures

A single lesion per patients was treated with HeberFERON®. In the case of patients with more than one lesion, the tumor to be treated was select as per size (with the largest diameter), or the lesion nearest to the others ones, o by the localization in high risk zone (mainly eyelids, periocular, in the ears, or nose).

The drug was administered by trained personnel, perilesional and intradermal, or intramuscularly, or subcutaneous, at a dose of 10.5 million international units (MIU) and with a frequency of three times a week for a minimum of three weeks in a total volume of 1 -5 mL. Treatment was performed on an outpatient basis in the appropriate hospital or polyclinic, medical office, or the patient’s house. Only the treated with HeberFERON^®^ lesions were evaluated for their characteristic (clinical subtype, size, localization, recurrence and aggressiveness) and clinical response to the treatment.

### Outcomes

The primary endpoint was the percentage of complete responses (CR) after one cycle of administration (3 weeks of treatment) and a follow for final response evaluation of 13 weeks from the end of treatment. The secondary endpoints were the demographic characteristics of patients and the characteristics of BCC lesions, according to the number of lesions.

### Statistical analysis

Quantitative variables were described with the arithmetic mean and its standard deviation and the median with its interquartile range. We used the absolute and relative frequency (%) for qualitative variables.

Influence of demographic and tumor variables on response were tested using univariate analyses by the chi-squared as association measure with the 95% confidence interval (CI) associated. The 95% CI were estimated for outcome variables. p<0.05 was considered statistically significant. Statistical analysis was performed using the Windows software package SPSS (version 25).

## Results

### Characteristic of patients

Out of 640 patients were recorder as treated with HeberFERON^®^ between 20 January 2017 and 11 December 2019 in the medical habitual practice in Cuba. Among them, 273 (42.7%) were females and 367 (57.3%) were males, with a male/female ratio of 1.34/1. The average age of evaluated patients was 67.9±13.8 years, with no differences between female (67.0±14.9 years) and males (68.6±12 years). Age ranged from 23 to 113 years, and most case (414; 68.7%) involved patients older than 65 years. The prevalent skin phototypes were type II (50.5%) and type III (32.5%), see Table 1 for details.

**Table 1.** Baseline characteristics and demographics of all patients

| Characteristics | Overall population (n=640) | One lesion (n=465) | ≥ 2 lesions (n=175) | 2-4 lesions (n=131) | ≥ 5 lesions (n=44) | p |
| --- | --- | --- | --- | --- | --- | --- |
| **Age,**<br>Mean (SD), years<br>Median (min, max), years | 67.9 ± 13.8<br>70 (28-113) | 67.7 ±13.8<br>70 (28-113) | 68.42 ±13.6<br>70 (29-94) | 70 ±12,7<br>72 (29-94) | 64.1 ±15.6<br>64.5 (33-92) | 0.048* |
| **Age male**<br>Mean (SD), years<br>Median (min, max), years | 68.6±12.9<br>70 (28-113) |  |  |  |  | 0.142* |
| **Age female** Mean (SD), years<br>Median (min, max), years | 67.0±14.9<br>70 (29-95) |  |  |  |  |  |
| **Sex, n (%)**<br>Male<br>Female | 367 (57.3%)<br>273 (42.7%) | 261 (56.1%)<br>204 (43.9%) | 106 (60.6%)<br>69 (39.4%) | 77 (58.8%)<br>54 (41.2%) | 29 (65.9%)<br>15 (34.1%) | 0.425 |
| **Skin photo type, n (%)**<br>Type I<br>Type II<br>Type III<br>Type IV<br>Type V | 60 (9.4%)<br>323 (50.5%)<br>225 (35.2%)<br>30 (4.7%)<br>2 (0.3%) | 41 (8.8 %)<br>234 (50.3 %)<br>163 (35.1 %)<br>25 (5.4 %)<br>2 (0.4 %) | 19 (10.9%)<br>89 (50.9%)<br>62 (35.4%)<br>5 (2.9%)<br>0 (0%) | 13 (9.9 %)<br>70 (53.4 %)<br>44 (33.6 %)<br>4 (3.1 %)<br>0 (0.0%) | 6 (13.6 %)<br>19 (43.2 %)<br>18 (40.9 %)<br>1 (2.3 %)<br>0 (0.0%) | NA |
| **Treated lesions size, n (%)**<br>Mean (SD), cm<br>Median (min-max), cm<br>≤ 2 cm<br>> 2 cm | 2.0 ±1.56<br>1.5 (0.1-12)<br>442 (69.1%)<br>198 (30.9%) | 2.0±1.4<br>1.7 (0.2-9.0)<br>319 (68.6%)<br>146 (31.4%) | 2.0±1.9<br>1.4 (0.1-12)<br>(70.3%)<br>52 (29.7%) | 2.2±2.04<br>1.5 (0.1-12)<br>88 (67.2%)<br>43 (32.8%) | 1.3±1.1<br>1 (0.4-4.7)<br>35 (79.5%)<br>9 (20.5%) | 0.015*<br>0.283<sup>s</sup> |
| **Type of the treated tumor, n (%)**<br>Primary tumor<br>Recurrent tumor | 407 (63.6%)<br>233 (36.4%) | 309 (66.5 %)<br>156 (33.5 %) | 98 (56.0%)<br>77 (44%) | 71 (54.2 %)<br>60 (45.8 %) | 27 (61.4 %)<br>17 (38.6 %) | 0.035 |
| **Clinical subtypes, n (%)** |  |  |  |  |  |  |
| Nodular | 290 (45.3%) | 216 (46.5%) | 74 (42.3%) | 58 (44.3%) | 16 (36.4%) | NA |
| Nodular ulcerative | 181 (28.3%) | 121 (26.0%) | 60 (34.3%) | 40 (30.5%) | 20 (45.5%) |  |
| Superficial | 88 (13.8%) | 65 (14.0%) | 23 (13.1%) | 19 (14.5%) | 4 (9.1%) |  |
| Nodular pigmented | 40 (6.3%) | 33 (7.1%) | 7 (0.4%) | 5 (3.8%) | 2 (4.5%) |  |
| Morpheaform | 25 (3.9%) | 19 (4.1%) | 6 (3.4%) | 6 (4.6%) | 0 (0.0%) |  |
| Infiltrative | 6 (0.9%) | 4 (0.9%) | 2 (1.1%) | 2 (1.5%) | 0 (0.0%) |  |
| Nodular cystic | 5 (0.8%) | 4 (0.9%) | 1 (0.6%) | 1 (0.8%) | 0 (0.0%) |  |
| Other | 5 (0.8%) | 3 (0.6%) | 2 (1.1%) | 0 (0.0%) | 2 (4.5%) |  |
| **Lesions sites, n (%)** |  |  |  |  |  |  |
| Nose | 232 (36.3%) | 184 (31.6%) | 48 (27.4%) | 41 (31.3%) | 7 (15.9%) | NA |
| Face | 99 (15.5%) | 55 (11.8%) | 44 (25.1%) | 30 (22.9%) | 14 (31.8%) |  |
| Ears | 72 (11.3%) | 53 (11.4%) | 19 (10.9%) | 16 (12.2%) | 3 (6.8%) |  |
| Trunk | 41 (6.4%) | 27 (5.8%) | 14 (8.0%) | 7 (5.3%) | 7 (15.9%) |  |
| Other | 41 (6.4%) | 34 (7.3%) | 7 (4.0%) | 5 (3.8%) | 2 (4.5%) |  |
| Cheek | 35 (5.5%) | 29 (6.2%) | 6 (3.4%) | 5 (3.8%) | 1 (2.3%) |  |
| Eyelids | 32 (5.0%) | 29 (6.2%) | 3 (1.7%) | 3 (2.3%) | 0 (0.0%) |  |
| Forehead | 31 (4.8%) | 19 (4.1%) | 12 (6.9%) | 9 (6.9%) | 3 (6.8%) |  |
| Eye internal canthus | 24 (3.8%) | 14 (3.0%) | 10 (5.7%) | 9 (6.9%) | 1 (2.3%) |  |
| Scalp | 11 (1.7%) | 6 (1.3%) | 5 (2.9%) | 3 (2.3%) | 2 (4.5%) |  |
| Neck | 6 (0.9%) | 3 (0.6%) | 3 (1.7%) | 3 (2.3%) | 0 (0.0%) |  |
| Upper limbs | 7 (1.0%) | 5 (1.0%) | 2 (1.1%) | 0 (0.0%) | 2 (4.5%) |  |
| Libs | 5 (0.8%) | 4(0.9%) | 1 (0.6%) | 0 (0.0%) | 1 (2.3%) |  |
| Eye external canthus | 4 (0.6%) | 3 (0.6%) | 1 (0.6%) | 0 (0.0%) | 1 (2.3%) |  |

NA: Non- applicable, ANOVA test (\*); Chi square test (\$).

The characteristics of the theses lesions, according to the number of lesions, are described in Table I. The average size of treated tumors was 2.0 ±1.56 cm; with a range from 0.1 to 12 cm. 30.9% of tumor measured more than 2 cm in larger diameter, and 31% were recurrent after surgery. The frequency of diagnosed clinical subtypes were nodular (45.3%), nodular ulcerative (28.3%), superficial (13.8%), nodular pigmented (6.3%), morpheaform (3.9%), infiltrative (0.9%) and nodular cystic (0.8%).

Topographic distribution (Table 1) of treated BCC shows the presence of tumors in nose (232; 36.3%), face (99; 15.5%), ears (72; 11.3%), trunk (41; 6.4%), cheek (35; 5.5%), eyelids (32;5.0%), forehead (31; 4.8%), eyes edges (28;4.4%), scalp (11;1.7%), limbs (7;1.0%), neck (6;0.9%), and libs (5;0.8%). Infrequent localizations were grouped as others (41; 6.4%).

Patients with single lesion (465; 72.6%) were the most abundant (Table 1), followed by those with 2 to 4 lesions (131; 20.5%) and patients with 5 or more lesions (44; 6.9%). Age differences between groups were found (p=0.048). The tumor size was smaller in the cohort of patients with more than 5 lesions (p=0.015). Primary tumors predominated in the group of one lesion (66.5%), and recurrent tumors were more represented among patients with 2-4 lesions (45.8%). Nodular BCC is more frequent in patients with single (46.5%) or 2-4 lesions (44.3%), whereas in a cohort with ≥ 5 lesions prevails the subtype nodular ulcerative (45.5%). Tumors were localized mainly in the nose with an overall distribution of 36.3%. However the face and the trunk were the zones of higher number of tumors for those patients with ≥ 5 lesions, 31.8% and 15.9%, respectively.

### HeberFERON effectivity

The results show clinical response distributed as follows: CR=61.9%, PR=32.7%, with an ORR of 94.2%. Disease stabilization of 5.8% was detected. Overall, the HeberFERON^®^ showed a 100% of disease control, with no progression reported (Table 2).

**Table 2.** Clinical response of BCC treated with HeberFERON.

| Characteristic | Clinical response |  |  |  | P/P<sub>ORR</sub> (chi) |
| --- | --- | --- | --- | --- | --- |
|  | CR, n (%) | PR, n (%) | SD, n (%) | ORR, n (%) |  |
| All patients | 394 (61.6%) | 209 (32.7%) | 37 (5.8%) | 603 (94.2%) | 0.000 |
| Sex |  |  |  |  |  |
| Male | 220 (60.0%) | 121 (33.0%) | 26 (7.1%) | 341 (93.0%) | 0.232/0.101 |
| Female | 174 (63.7%) | 88 (32.2%) | 11 (4.0%) | 262 (96.0%) |  |
| Age |  |  |  |  |  |
| < 65 | 151 (66.8%) | 66 (29.2%) | 9 (4.0%) | 217 (96.0%) | 0.091/0.150 |
| ≥ 65 | 243 (58.7%) | 143 (34.5%) | 28 (6.8%) | 286 (93.2%) |  |
| Skin phototype |  |  |  |  |  |
| Type I | 41 (68.3%) | 17 (28.3%) | 2 (3.3%) | 58 (96.7%) | NA |
| Type II | 201 (62.2%) | 96 (29.7%) | 26 (8.0%) | 297 (92.0%) |  |
| Type III | 129 (57.3%) | 88 (39.1%) | 8 (3.6%) | 317 (96.4%) |  |
| Type IV | 21 (70.0%) | 8 (26.7%) | 1 (3.3%) | 29 (96.7%) |  |
| Type V | 2 (100%) | 0 (0.0%) | 0 (0.0%) | 2 (100%) |  |
| Type of tumor |  |  |  |  |  |
| Primary | 254 (62.4%) | 136 (36.4%) | 17 (4.2%) | 390 (95.8%) | 0.071/0.22 |
| Recurrent | 140 (60.1%) | 73 (31.3%) | 20 (8.6%) | 213 (91.4%) |  |
| Tumor size |  |  |  |  |  |
| ≤ 2 cm | 307 (69.5%) | 122 (27.6%) | 13 (2.9%) | 429 (97.1%) | 0.000/0.000 |
| > 2 cm | 87 (43.9%) | 87 (43.9%) | 24 (12.1%) | 174 (87.9%) |  |
| Clinical subtype |  |  |  |  |  |
| Nodular | 186 (64.15) | 90 (31.0%) | 14 (4.8%) | 276 (95.6%) | NA |
| Nodular ulcerative | 100 (55.2%) | 66 (36.5%) | 15 (8.3%) | 166 (91.7%) |  |
| Superficial | 57 (64.8%) | 28 (31.8%) | 3 (3.4%) | 85 (96.6%) |  |
| Nodular pigmented | 23 (57.5%) | 15 (37.5%) | 2 (5.0%) | 38 (95.0%) |  |
| Morpheaform | 18 (72.0%) | 6 (24.0%) | 1 (4.0%) | 24 (96.0%) |  |
| Infiltrative | 4 (66.7%) | 2 (33.3%) | 0 (0.0%) | 6 (100%) |  |
| Nodular cystic | 3 (60.0%) | 0 (0.0%) | 2 (40.0%) | 3 (60%) |  |
| Other | 3 (60.0%) | 2 (40.0%) | 0 (0.0%) | 5 (100%) |  |

**CR:** Complete response, **PR:** Partial response, **ORR:** Objective response rate, **SD:** Stable disease, **DC:** Disease control

The best responder subtypes of BCC to HeberFERON^®^ were the more aggressive tumors, morpheaform with CR of 72% (ORR=96%), followed by the infiltrative with 66.7% (ORR=100%). A good responder were superficial (CR=64.8%, ORR=96.6%) and nodular (CR=64.15%, ORR=95.6%) BCC. These differences were significant for the ORR.

The clinical response rate by the number of tumor per patient (Table 3) showed that in a cohort of patients with a single tumor the CR is significant higher (67.5%, p=0.000) with respect to the group of 2-4 lesions (51%) or ≥ 5 lesions (30%). In several patients with more than one BCC the route of HeberFERON^®^ administration was parentally, intramuscular or subcutaneous. Analyzing the clinical response rate with respect to the route of administration (Table 4) is noted that the most effective route, with a tendency to significance, was the perilesional/intradermic way of administration with 63.5% of CR (Table 4).

**Table 3.** Clinical response of BCC treated with HeberFERON by number of lesions.

| Response | Groups by number of lesions |  |  |  |
| --- | --- | --- | --- | --- |
|  | One lesion<br>(n=465) | ≥ 2 lesions<br>(n=175) | 2-4 lesions<br>(n=131) | ≥ 5 lesions<br>(n=44) |
| CR, n (%) | 314 (67.5%) | 80 (45.7%) | 67 (51.0%) | 13 (29.5%) |
| PR, n (%) | 132 (28.4%) | 77 (44.0 %) | 51 (38.9%) | 26 (59.1%) |
| SD, n (%) | 19 (4.1%) | 18 (10.3%) | 13 (9.9%) | 5 (11.4%) |
| p | 0.000 |  | 0.000 |  |
| ORR, n (%) | 446 (95.9%) | 157 (89.7%) | 118 (90.1%) | 39 (88.6%) |
| p | 0.04 |  | 0.011 |  |

**Table 4.** Response rate to HeberFERON by administration route.

| Response | Routes of HeberFERON administration |  |  |  |
| --- | --- | --- | --- | --- |
|  | Intradermic<br>(N=575) | Intramuscular<br>(N=47) | Subcutaneous<br>(N=18) | p |
| CR, n (%) | 365 (63.5%) | 19 (40.4%) | 10 (55.6%) | 0.07 |
| PR, n (%) | 175 (30.4%) | 26 (55.3%) | 8 (3.8%) |  |
| ORR, n (%) | 540 (93.9%) | 45 (95.7%) | 10 (100%) |  |

Summarizing the results, we found that the nose was the region of higher frequency of tumors and the nodular clinical subtype was the predominant. We have detected that the HeberFERON^®^ was highly effective in BCC in real world conditions, with the aggressive tumors subtypes more sensible to the anti-proliferative effect of these combination of IFNs. Tumor with larger size and patients with ≥ 5 tumors had lesser response to the anti-tumor effect of HeberFERON^®^. Perilesional/intradermic administration of IFNs resulted with the best clinical response rate.

## DISCUSIÓN

BCC is the most common type of human tumor. Patients with BCC have a 17-fold increased risk of a new BCC compared to the general population, followed by a 3-fold increase in the risk of squamous cell carcinoma and a two-fold risk of developing melanoma and other cancers related to radiations^25,6^. Approximately 80% of BCCs occur in the head and neck region with major impact in health-related quality of life of the patient as a consequence of disfiguring skin changes^26^ and the interventions applied^27,28^.

The analysis of data from the NCEP-HFN since 2017 to 2019 showed that the Cuban population with a diagnosis of BCC, including those with multiple lesions (27.3%), is a population of a median age of 70 years, with more aged patients (median 72 years) bearing more than two BCC. The highest incidence of the disease occurred in patients with Type II (50.5%), and Type III skin phototypes (32.5%), predominantly male (57.3%).

In a clinical and histopathological study carried out in Sinaloa, Mexico^29^, it is reported that BCC prevails in the population between 60 and 70 years of age, probably due to the accumulation of sun damage over time^30,31^. In this study the female sex predominated in contrast with the prevalent literature data that describes the higher frequency of BCC in men, likely associated with lifestyles, weather conditions and occupations, which make the development of BCC in females more frequent^32^.

Many patients are prone to develop multiple primaries BCC, with reports showing 12% to 46% of subjects with more than a single skin lesion.^33^ In this study men showed propensity to develop multiple BCC in concordance with data of other reports^34,35,36^. Further, in the subgroups of patients with multiple BCC, the evaluated tumors developed more frequently in nose and face. The results did not reproduce the observations of other authors that found more multiple BCC in trunk (back) and upper limbs and less in face^37, 33^. However in the case of patients with ≥ 5 lesions, our study detected more frequency of tumor in the trunk and face.

We found 84% treated BCC located in the head, a higher frequency with respect to Villani et al.^38^ that studying advanced or metastatic BCC in real world conditions described 62.5% of BCC located in the head and neck.

Likewise, we identified as clinical more common BCC subtypes nodular, nodular ulcerative and superficial, similar as reported by Villani et al.^38^ showing more frequency for ulcerative (45.8%), multiple nodular and superficial (29.2%) and nodular (20.8%) clinical variants of BCC. In our study in subjects with ≥ 5 tumors the prevalent clinical subtype was nodular ulcerative. Conversely, Bartos et al. ^33^ described as more common the superficial BCC and less frequent nodular and infiltrative subtypes. Similar as reported by others^39^ no influence of skin photo type was found on the development of a second and more skin BCCs.

The development of therapies to enhance the action of components of immunity on BCC and specifically, the use of IFNs has been investigated for several years. IFNs mediate antitumor effects either indirectly by modulating the immune and anti-angiogenic response or directly, affecting the proliferation and differentiation of tumor cells^40^.

We are reporting for the first time a real world data of the use of the combination of IFNs in patients with single or multiple BCC. There are no literature references of effectivity with the use the combination of IFNs in these cohorts of patients. The high overall response rate of 96% (CR=61.6%) has not been reported never before for BCC with the use of IFNs or other non-surgical approaches in BCC in real-life practice.

The infiltrative and morpheaform cases of BCC are more aggressive, characterized by a diffuse and destructive growth pattern^41, 42^. Interesting, the HeberFERON^®^ treatment was likely more effective in these two clinical subtypes.

When performing these analyzes, subdividing the patients by number of lesions, we found similar clinical ORR in the three groups. However, lower percentage of CR was found in the group of patients with multiple BCC, with the lowest rate in the cohort of patients with ≥ 5 lesions.

BCC is an immunogenic tumor^43,44^. Evidences highlighted the critical role of immune surveillance in the control of BCC^45,46^. This lower clinical response in patient with multiple BCC treated with the combinations of IFNs could be associated with an immune compromise^47^ or tumor microenvironment subversion^48^ characterized by a deficient antigen presentation process (lack of HLA-I functionality, low immune stimulatory effects and prevalence of regulatory immune cell tumor microenvironment).

In the context of resistance of skin tumor to inhibitors of hedgehog signaling and immune check point inhibitor^49^, the combination of IFNs alpha and IFN gamma appears as a plausible therapeutic option for wide number of BCC, that may include intermediate and high risk, multiple or advanced BCC.

The inclusion of IFN gamma in this formulation could reinforce the immune regulatory effect of IFNs and contribute to a more effective immune-mediated antitumor effect. The role of IFN gamma in restoring tumor antigen presentation and immune check point expression has been expected^50, 51^.

Surgery is the most frequently applied BCC treatment, but nonsurgical modalities do also have an essential role in clinical practice^52^. Further studies are required to confirm the fundamental role of the combination of IFN alpha and gamma in the effective control of BCC tumor growth in the real world environment.

## Data Availability

Qualified individuals may request access to the de-identified participant data, anonymized clinical study reports, informed consent forms, through submission of a proposal with a defined research question to the corresponding author, Bello-Rivero Iraldo, provided that the necessary data protection and ethical committee approvals are in compliance with the trial. An agreement for transfer of these data will be required.

## Declarations

Study execution conformed to the ethical principles of the Declaration of Helsinki and the International Council for Harmonization of Good Clinical Practice guidelines. The authors were responsible for designing the trial and for collecting and analyzing the data. The following ethical and/or research institutional committee waived the approval of the study in their respectively institutions in Cuba.

1. Jose Avila Serrano Polyclinic Ethical Committee waived ethical oversight of the study. Municipality of Gibara, Holguin Province.
2. Center Polyclinic Ethical Committee waived ethical oversight of the study. Santi Spiritus City, Santi Spiritus Province.
3. Antonio Luaces Iraola Hospital Ethical and Research Committee waived ethical oversight of the study. Ciego de Avila City, Ciego de Avila Province.
4. Faustino Perez Hospital Ethical Committee waived ethical oversight of the study. Matanzas City, Matanzas Province.
5. Carlos J. Finlay Polyclinic Ethical Committee waived ethical oversight of the study Municipality of Colon, Matanzas Province.
6. Arnaldo Milian Castro Hospital Ethical and Research Committee waived ethical oversight of the study. Villa Clara City; Villa Clara Province.
7. Maria Curi Oncological Hospital and Amalia Simoni Hospital Ethical and Research Committees waived ethical oversight of the study. Camaguey City, Camaguey Province.
8. Gustavo Aldereguia Hospital, Ethical and Research Committee waived ethical oversight of the study. Cienfuegos City, Cienfuegos Province.
9. Pablo Noriega Polyclinic Ethical Committee waived ethical oversight of the study. Mayabeque Province.
10. Comandante Pinares Hospital Ethical and Research Committee waived ethical oversight of the study, Municipality of San Cristobal, Artemisa Province.
11. Heroes del Baire Hospital Ethical Committee waived ethical oversight of the study. Nueva Gerona City, Special Municipality Island of Youth, Cuba.
12. Celia Sanchez Manduley Hospital Ethical Committee waived ethical oversight of the study, Manzanillo City, Granma Province.
13. Juan Bruno Zayas Hospital Ethical and Research Committee waived ethical oversight of the study, Santiago de Cuba City, Santiago de Cuba Province.
14. Ernesto Guevara Hospital Ethical and Research Committee waived ethical oversight of the study. Las Tunas City, Las Tunas Province.
15. Leon Cuervo Rubio Hospital Ethical and Research Committee waived ethical oversight of the study. Pinar del Rio City, Pinar del Rio Province.
16. Agustinho Neto Hospital Ethical Committee waived ethical oversight of the study. Guantánamo City, Guantanamo Province.
17. Calixto Garcia Hospital Ethical and Research Committee waived ethical oversight of the study. Havana City, Havana Province.
18. Center of Medical Surgical Investigations Ethical and Research Committee waived ethical oversight of the study. Havana City, Havana Province.
19. Salvador Allende Hospital Ethical and Research Committee waived ethical oversight of the study. Havana City, Havana Province.
20. Manuel Fajardo Hospital Ethical and Research Committee waived ethical oversight of the study. Havana City, Havana Province.

## Authors’ contributions

JVA, VSL, DSD, YPR, MFM, ABP, MLJ, YBC, RMP, MCA, AMA, RMB, JMP, LPG, NLP, YRH, MRT, IFR, APL, YLG, SSCh, YLA, were the clinical investigators [health provincial or hospital coordinators of the NCEP-HFN that prescribed and supervised the treatment of patients in the clinical practice, and recorded the information in medical records of the patients included from the clinical site in their provinces. YRA and ORA, coordinated, executed and supervised the NCEP-HFN at the health institutions in Cuba. RHR was responsible for data base and its quality. YGV was responsible for statistical analyses, clinical results interpretation, revision and manuscript editing. MHC contributed to the design of trial protocol. CMS contributed to the design of trial protocol, data analysis and contributing to writing of the manuscript. YDR contributed to the design of trial protocol. IBR designed and wrote the protocol, supervised the trial, interpreted and discussed all the data and results, and contributed to the writing of the manuscript. All authors read and approved the final manuscript. All authors read and approved the final manuscript.

## Acknowledgements

We thank the specialists and technical staff from the Dermatological Services/Department from the health institution participating in the of NCEP-HFN and in the study. We also acknowledge the supervision of health coordinators of CIGB in their supervision of the execution of NCEP-HFN that guarantied the rapid and secure collection of the patient’s data and clinical outcomes.

